# Portable sequencing in the field and the classroom: a retrospective examination of the circulation of DENV1 and DENV2 in Brazil

**DOI:** 10.1101/2020.09.01.20183301

**Authors:** Talita Émile Ribeiro Adelino, Marta Giovanetti, Vagner Fonseca, Joilson Xavier, Álvaro Salgado de Abreu, Valdinete Alves do Nascimento, Luiz Henrique Ferraz Demarchi, Marluce Aparecida Assunção Oliveira, Vinícius Lemes da Silva, Arabela Leal e Silva de Mello, Gabriel Muricy Cunha, Roselene Hans Santos, Elaine Cristina de Oliveira, Jorge Antônio Chamon Júnior, Felipe Campos de Melo Iani, Ana Maria Bispo de Filippis, André Luiz de Abreu, Ronaldo de Jesus, Carlos Frederico Campelo de Albuquerque, Jairo Mendez Rico, Rodrigo Fabiano do Carmo Said, Joscélio Aguiar Silva, Noely Fabiana Oliveira de Moura, Priscila Leite, Lívia Carla Vinhal Frutuoso, Simone Kashima Haddad, Alexander Martínez, Fernanda Khouri Barreto, Cynthia Carolina Vazquez, Rivaldo Venâncio da Cunha, Emerson Luiz Lima Araújo, Stephane Fraga de Oliveira Tosta, Allison de Araújo Fabri, Flávia Löwen Levy Chalhoub, Poliana da Silva Lemos, Fernanda de Bruycker-Nogueira, Gislene Garcia de Castro Lichs, Marina Castilhos Souza Umaki Zardin, Fátima María Cardozo Segovia, Crhistinne Cavalheiro Maymone Gonçalves, Zoraida Del Carmen Fernandez Grillo, Svetoslav Nanev Slavov, Luiz Augusto Pereira, Ana Flávia Mendonça, Felicidade Mota Pereira, Jurandy Júnior Ferraz de Magalhães, Agenor de Castro Moreira dos Santos, Maricélia Maia de Lima, Rita Maria Ribeiro Nogueira, Aristóteles Goes Neto, Vasco Ariston de Carvalho Azevedo, Dario Brock Ramalho, Wanderson Kleber Oliveira, Arnaldo Correia de Medeiros, Victor Pimentel, Latin American Genomic Surveillance Arboviral Network, Edward C Holmes, Tulio de Oliveira, José Lourenço, Luiz Carlos Junior Alcantara

## Abstract

**Background:** Brazil has experienced a large dengue virus (DENV) epidemic in 2019, highlighting a continual struggle with effective control and public health preparedness. Brazil is a world leader in real-time genomic surveillance of arboviruses, although such technology and expertise remains inaccessible for the vast majority of local researchers and public health workers. In 2019, we led field and classroom initiatives for the genomic surveillance of DENV in Brazil.

**Methods:** Oxford Nanopore MinION technology was used for sequencing, focusing on generating DENV1 and DENV2 complete genomes. Using phylogenetic and epidemiological approaches conducted in real-time during a training program and subsequently through online channels, we explored the recent spatio-temporal evolution and spread of these viruses in Brazil.

**Findings:** In the years following the Zika virus epidemic (2017-2018) reporting was at an all-time low, and significant increases in reported cases and deaths in 2019 did not reflect a higher case fatality ratio. Estimated transmission potential and reporting of other arboviruses suggests that neither arboviral reporting saturation nor climatic factors can easily explain the post-Zika period and resurgence in 2019 (respectively). Phylogenetic analysis revealed complex patterns of transmission, with lineage co-circulation and replacement, in which the North and the Southeast acted as sources of dispersion to other regions. We identified two lineages within the already reported DENV2 BR-4 clade, for which the effective reproduction number had seasonal signatures alike reported cases, with a temporal increase towards 2019 mirroring the large epidemic that year.

**Interpretation:** We describe the recent evolution and diffusion of DENV1 and DENV2 in Brazil. Importantly, the surveillance outputs and training initiative here described serve as proof-of-concept of the potential of portable sequencing for both research and local capacity building in the area of genomic surveillance of arboviruses.

**Funding:** Decit, SCTIE, BrMoH, CNPq, CAPES, EU Horizon 2020 through ZIKAlliance and STARBIOS2.

## RESEARCH IN CONTEXT

### Evidence before this study

Since the early 1980s a varying number of DENV serotypes have been endemic in Brazil, with the country reporting the highest number of cases worldwide, at approximately 16 million infections. Despite such hyperendemicity, there is still relatively limited information on the evolution, transmission and spread of DENV within Brazil, which is essential for epidemic preparedness, response and control. Previous studies have described complex transmission dynamics in Brazil, in which DENV persists in a constant spatio-temporal flux, with the hyperendemic co-circulation and changing frequencies of multiple serotypes fueled by the Caribbean region and northern Brazilian states. The country has faced several severe mosquito-borne epidemics in the past five years, but still faces significant challenges in outbreak preparedness and control. As such, there is much to be gained in Brazil from dedicated training programs in the area of genomic surveillance with a focus on laboratory and computational methodologies that target postgraduate students, laboratory technicians and health practitioners.

### Added value of this study

Using a combined strategy based on two initiatives – field genomic surveillance with a mobile laboratory in the Brazilian Midwest, and a classroom genomic surveillance training program targeting universities and public health laboratories networks in the Americas – we generated 227 novel genome sequences of DENV1 and DENV2 from 85 municipalities across Brazil (2015-2019), equating to +50% increase in the number of currently available Brazilian genomes in public databases. Using both phylogenetic and epidemiological models we retrospectively reconstruct and describe the recent transmission history of DENV1 and DENV2 in Brazil. We find mosquito-viral suitability and the effective reproduction number dynamics to reflect and explain much of the temporal variation in reported cases. We also describe instances of lineage and serotype replacement through time, as well as support for the role of the Caribbean, Northern and Southeastern Brazilian states in viral dissemination.

### Implications of all the available evidence

Our findings highlight the important role of the Caribbean, the Northern and Southeastern states of Brazil in viral dissemination, supporting the notion that active surveillance in these regions is essential for virus control both in Brazil and nearby regions. Our results are also consistent with ecological suitability of *Aedes* spp. and insufficient human herd immunity in driving the timing and size of local dengue epidemics in Brazil. Since most of the new genomic data was generated and analysed in real-time within the genomic surveillance training program, we present a proof-of-concept that portable sequencing technologies offer unique opportunities for capacity building in settings where post-graduate students, laboratory technicians and health practitioners do not have institutionalized opportunities to develop expertise in the area of genomic surveillance.

## INTRODUCTION

Dengue virus (DENV) has spread extensively over the past decade and now poses a threat to about one-third of the global human population, primarily inhabitants of tropical and subtropical regions where mosquito vectors of the genus *Aedes* are widely distributed.^1^ Increases in human mobility and population growth, unplanned urbanization, globalization, climate change, and unsuccessful vector control programs have contributed to DENV’s expansion, making it a major public health threat at a global scale.^2^ Infection with the virus causes a wide spectrum of clinical manifestations, including asymptomatic or mild self-limiting disease (Dengue Fever), or severe disease characterized by vascular leakage and haemorrhagic symptoms, such as Dengue haemorrhagic Fever (DHF) and Dengue Shock Syndrome (DSS).^3^ DENV is a single-stranded, positive-sense RNA virus with a genome of ~11,000 kb belonging to the family *Flaviviridae* (genus *Flavivirus)*. Its genome encodes a polyprotein that is post-translationally processed into three structural (capsid, pre-membrane or membrane, and envelope) and seven non-structural (NS1, NS2a, NS2b, NS3, NS4a, NS4b, and NS5) proteins.^4^ DENV is classified into four antigenically distinct and genetically related serotypes (DENV 1-4), each of which contain several major clades (termed genotypes) that often have differing spatio-temporal distributions.^5^ DENV was successfully eradicated in many regions of South America during the mid-twentieth century,^6^ with resurgence first reported in Brazil in the early 1980s in the northern state of Roraima.^7^ Subsequently, the southern state of Rio de Janeiro played a pivotal role in the sequential resurgence of each serotype: DENV1 was reported to have been introduced there in 1986,^8^ DENV2 in 1990 simultaneously with the first epidemic of DHF,^9^ DENV3 in 2000,^10^ and finally DENV4 in 2010.^11^ Brazil can now be considered hyperendemic for dengue, reporting the highest number of dengue cases worldwide, with approximately 16 million notified infections since the 1980s.^12^

Different serotypes have caused unexpected large epidemics in Brazil over the past 20 years, with particularly problematic outbreaks in 2002, 2008, 2010, and 2012-2013.^13^ In the years following the Zika virus epidemic (2017-2018) dengue reporting was surprisingly low.^13^ However, the re-emergence of DENV2 in 2019 followed a report of a staggering 1,544,987 probable cases and 782 confirmed deaths (until epidemiological week – EW52 2019).^14^ Even with a long history of DENV circulation and recent experience with large outbreaks of other arboviruses (e.g. Zika, chikungunya, yellow fever), Brazil continues to struggle with effective mosquito control and public health preparedness.^15^ In contrast, over the last five years, the country has become a world leader in real-time genomic surveillance of arboviruses, contributing greatly to our current understanding of the molecular evolution, spread, and persistence of such viruses (e.g. https://www.zibraproject.org and https://www.zibra2project.org). Despite this, expertise on the methodologies involved with real-time genomic surveillance remains inaccessible to the vast majority of local researchers and public health workers.^16^ There is much to be gained in Brazil from dedicated training programs in the area of genomic surveillance, targeting those who do not have institutionalized access to expertise in this area.

Employing genomic surveillance in the field and in the classroom under a large training program, we generated and analysed 227 novel complete genome sequences of DENV1 and DENV2. Informed by a mix of public and newly generated genomic data, climate, and epidemiological time series, we use phylogenetic and epidemiological models to reconstruct the recent transmission history of these serotypes in Brazil. We find a complex dynamic history that vastly supports previously proposed events of viral movement, replacement, and co-circulation. By generating and analyzing most of the data in real-time within the training program attended by participants of various educational and professional backgrounds from across the Americas, we further provide a proof-of-concept of the unique opportunities that portable sequencing technologies offer for local capacity building.

## MATERIALS AND METHODS

### Ethics statement

This project was reviewed and approved by the Comissão Nacional de Ética em Pesquisa (CONEP) from the Brazilian Ministry of Health (BrMoH), as part of the arboviral genomic surveillance efforts within the terms of Resolution 510/2016 of CONEP, by the Pan American Health Organization Ethics Review Committee (PAHOERC) (Ref. No. PAHO-2016-08-0029), and by the Oswaldo Cruz Foundation Ethics Committee (CAAE: 90249218.6.1001.5248). The samples processed in this study were obtained anonymously from material exceeding the routine diagnosis of arboviruses in Brazilian public health laboratories that belong to the BrMoH’s public network.

### Field genomic surveillance with a mobile laboratory

In May 2019 we implemented an arbovirus surveillance project that took place across the Midwest of Brazil using a mobile genomics laboratory (Figure S1). This Brazilian-driven initiative, known as the ZiBRA-2 project, was supported by the BrMoH (https://www.zibra2project.org).

### Classroom genomic surveillance in a training program

In August 2019 a genomic surveillance training program organized by PAHO and BrMoH took place in Belo Horizonte (Minas Gerais state) under the title *“Nanopore-based genome sequencing technology for temporal investigation and epidemiology of dengue outbreak: training, research, surveillance, and scientific dissemination “*. The syllabus included practical and theoretical courses on a variety of subjects related to arbovirus research and surveillance, including mobile sequencing technologies, bioinformatics, phylogenetics, epidemiological modelling, and field epidemiology and entomology. The event targeted post-graduate students, laboratory technicians, and health practitioners in universities and laboratories across the Americas and was based on the principles of Responsible Research and Innovation (RRI).^17^ Details on the program can be found in Supplementary Text File 1.

### Sample collection and molecular diagnostic assays

Clinical samples from patients with suspected DENV infection were obtained for routine diagnostic purposes at local health services in different Brazilian municipalities. These samples were sent for molecular diagnosis to the respective, local Central Laboratory of Public Health (LACEN) from the Brazilian Federal District (DF) and from the states of Bahia (BA), Goiás (GO), Mato Grosso (MT), Mato Grosso do Sul (MS), Minas Gerais (MG), Pernambuco (PE), and Rio de Janeiro (RJ), according to the protocol established by the BrMoH. Samples processed from the state of São Paulo (SP) were collected by the Blood Center of Ribeirão Preto from volunteer blood donors eligible for blood donation and who reported adverse effects up to 14 days after donation.

Viral RNA was extracted from all clinical samples using the QIAmp Viral RNA Mini Kit (Qiagen) and tested by RT-qPCR for detection of DENV1-4. Selected samples with previous positive diagnostic results for DENV1-2 were processed in two steps: (1) 73 samples from the states of GO, MS, and MT were processed during the ZiBRA-2 project, (2) 175 samples from the DF and BA, GO, MG, PE, RJ, and SP were processed during the training program (both initiatives described in the section above). Samples from the 2019 outbreak, as well as available samples from previous epidemic waves in 2008 and between 2015-2018, were included for diagnostic screening.

### cDNA synthesis and whole genome sequencing in MinION

Samples were selected for sequencing based on the Ct value (≤35) and availability of epidemiological metadata, such as date of symptom onset, date of sample collection, sex, age, municipality of residence, symptoms, and disease classification. For complementary DNA synthesis, the SuperScript IV Reverse Transcriptase kit (Invitrogen) was used following the manufacturer’s instructions. The cDNA generated was subjected to sequencing multiplex PCR (35-cycles) using Q5 High Fidelity Hot-Start DNA Polymerase (NEB) and a set of specific primers designed by the CADDE project (https://www.caddecentre.org/) for sequencing the complete genomes of DENV1-2.

Amplicons were purified using 1x AMPure XP Beads (Beckman Coulter) and quantified on a Qubit 3.0 fluorimeter (Thermofisher Scientific) using Qubit™ dsDNA HS Assay Kit (Thermofisher Scientific). Of the 248 samples, 227 contained sufficient DNA (≥2ng/^L) to proceed to library preparation. DNA library preparation was performed using the Ligation Sequencing Kit (Oxford Nanopore Technologies) and the Native Barcoding Kit (NBD103, Oxford Nanopore Technologies).^18^ Sequencing libraries were generated from the barcoded products using the Genomic DNA Sequencing Kit SQK-MAP007/SQK-LSK208 (Oxford Nanopore Technologies) and loaded into a R9.4 flow cell (Oxford Nanopore Technologies).

### Generation of consensus sequences

Raw files were basecalled using Guppy and barcode demultiplexing was performed using qcat. Consensus sequences were generated by *de novo* assembling using Genome Detective (https://www.genomedetective.com/).^19^

### Phylogenetic analysis

DENV genotyping was performed using the Dengue Virus Typing Tool (https://www.genomedetective.com/app/typingtool/dengue/).^20^ To investigate the evolution and population dynamics of DENV1-2 in different regions, the DENV1 (n=57) and DENV2 (n=170) complete genome sequences generated in this study were combined with publicly available complete genome sequences from DENV1 genotype V (DENV1-V) and DENV2 genotype III (DENV2-III) as these represent the dominant genotypes in the Americas. The latter were retrieved from NCBI up to November 2019. Sequences without sampling date and location were excluded, as were sequences covering less than 50% of the viral genome.

Sequence alignment was performed using MAFFT^21^ and manually curated to remove artifacts using Aliview.^22^ Maximum Likelihood (ML) phylogenetic trees were estimated using IQ-TREE^23^ under the best-fit substitution model indicated by the Model Finder application implemented in IQ-TREE. Robustness of the tree topology was determined using 1,000 bootstrap replicates, and the presence of temporal signal was evaluated in TempEst^24^ through a regression of root-to-tip genetic distances against sampling time. Time-scaled phylogenetic trees were inferred using the BEAST package.^25^ The uncorrelated relaxed molecular clock model was chosen as indicated by estimating marginal likelihoods, also employing the codon based SRD06 model of nucleotide substitution and the non-parametric Bayesian Skyline coalescent model. A discrete phylogeographic model^26^ was used to reconstruct the virus spatial diffusion across the sampling locations. Discrete locations were initially defined as the country of sampling. However, a different resolution was applied according to sampling availability. Phylogeographic analyses were then performed by applying an asymmetric model of location transitioning coupled with the Bayesian Stochastic Search Variable Selection (BSSVS) procedure. Markov Chain Monte Carlo (MCMC) were run for sufficiently long to ensure stationarity and an adequate Effective Sample Size (ESS) of >200.

### Epidemiological data and integration with genomic data

Data from 2015 to 2019 of weekly notified and laboratory confirmed cases of infection by DENV in Brazil, as well as monthly fatal cases with confirmed dengue infection, were supplied by the BrMoH. A mosquito-viral suitability measure (index P) was estimated using the MVSE R-package.^27^ The index P measures the reproductive (transmission) potential of a single adult female mosquito in a completely susceptible host population and is informed by local temperature and humidity time trends. We used daily climatic data from the three largest cities of each macro region for which new sequences were generated (Midwest: Goiânia, Brasília, Campo Grande; Southeast: São Paulo, Rio de Janeiro, Belo Horizonte; Northeast: Salvador, Recife, Fortaleza), with data obtained from openweathermap.org.

### Estimating R_e_ from genetic sequences

We used birth-death models implemented in BEASTv2.4^28^ to estimate the effective reproduction number (R_e_). In this model, each infection may transmit at a rate λ and will become non-infectious at a rate *δ*. Upon becoming infected, each individual is sampled with a probability *s^s^*. The model enables the piecewise estimation of R_e_, *δ*, and *s* through time. We assumed sampling proportion *s* to be constant over time. Relaxing this assumption to allow the parameter *s* to be zero for the periods when no sequence data was available resulted in similar trends for R_e_, with wider Bayesian credible intervals. The rate *δ* was modelled using a lognormal prior with a mean of 14 days and a standard deviation of 0·5, which roughly corresponds to the sum of the intrinsic and extrinsic incubation period of dengue virus. The birth-death skyline (BDSKY) analysis was run for an independent MCMC chains of >100 steps, with parameters and trees being sampled once every 10,000 steps. After removal of 10% burn in, sampled parameters were combined using LogCombiner.

## RESULTS

A total of 248 RT-qPCR positive samples for DENV1 (n=62) and DENV2 (n=186) were screened. Of the samples tested, 227 (DENV1=57, DENV2=170) contained sufficient DNA (≥2ng/μL) to proceed to library preparation. For those positive samples, PCR cycle threshold (Ct) values were on average 22 (range: 15 to 33) and 23 (range: 16 to 33) for DENV1 and DENV2, respectively. Epidemiological details of the processed samples are provided in Table S1. To investigate the evolutionary history and transmission dynamics of DENV1 and DENV2, we used a portable nanopore sequencing approach^13^ to generate the complete genomes sequences from the 227 viral samples. The resulting average coverage was 89·3% for DENV1 and 87·6% for DENV2. Sequencing statistics are detailed in Table S2.

DENV1 sequences covered the time period August 2015 to July 2019 in DF (n=2) and five Brazilian states (BA=19; GO=10; MG=11; PE=11; SP=4), while DENV2 samples were collected between January 2016 and August 2019 in DF (n=1) and eight Brazilian states (BA=20; GO=31; MG=47; MS=59; MT=2; PE=3; RJ=1; SP=3) (Figure 1 panel A). Three older DENV2 samples from the GO state, sampled during 2008, were also included. The median age of patients was 28 years (range: 4 to 78 years of age) for DENV1 and 37 years (range: 4 to 90 years of age) for DENV2 cases. Considering the clinical outcome, 77·2% of DENV1 (44/57) and 81·2% of DENV2 (138/170) samples were obtained from patients without alarming clinical signs (DF), 3·5% of DENV1 (2/57) and 2·3% of DENV2 (4/170) corresponded to patients with severe dengue (DHF/DSS). Finally, 19·3% of DENV1 (11/57) and 16·5% of DENV2 (28/170) were obtained from fatal cases (Table S1).

**Figure 1.**
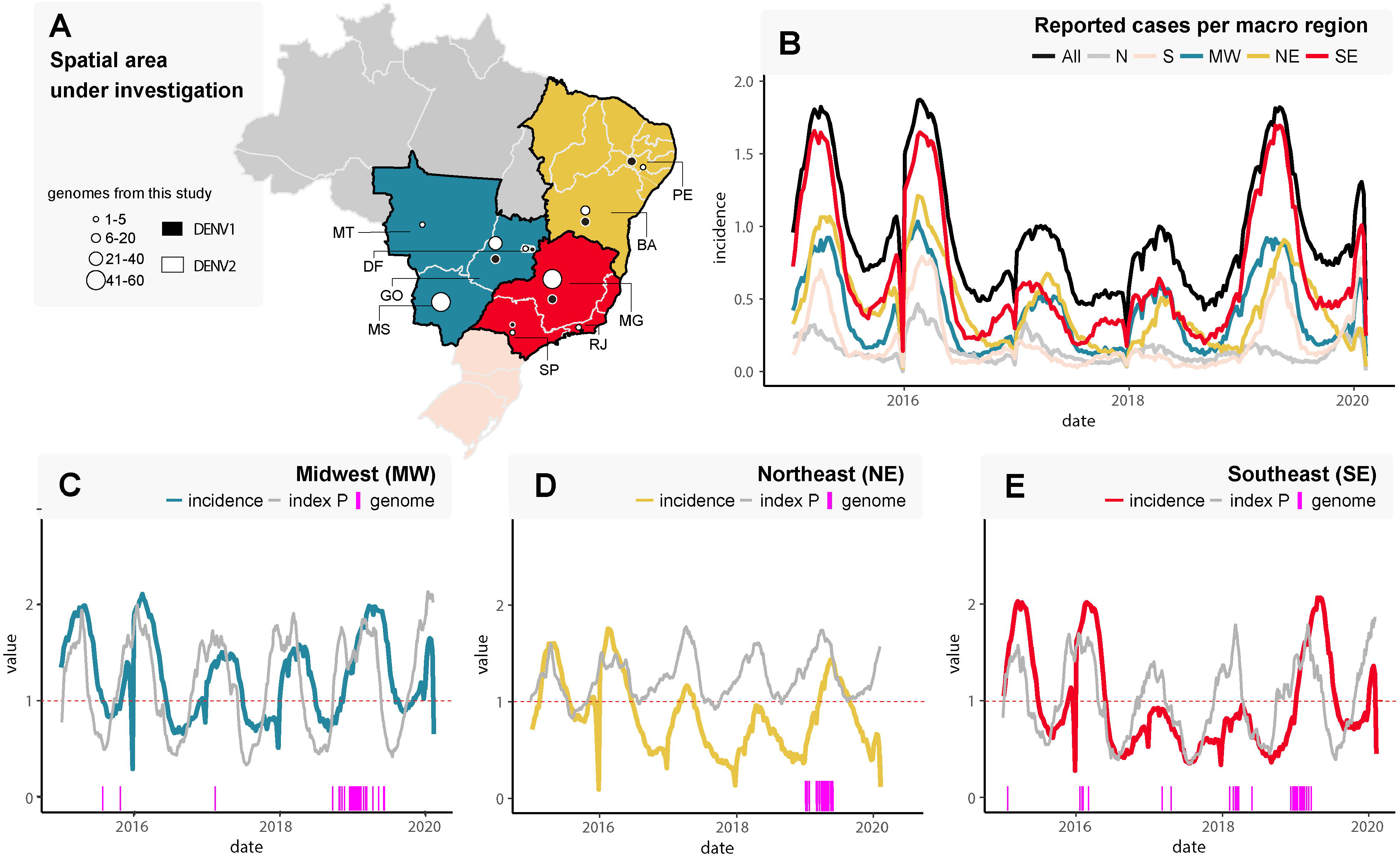
Spatial and temporal distribution of reported dengue cases in Brazil, 2015-2020. **A**. Map of Brazil showing the number of DENV1 and DENV2 new sequences by region and state. DF=Brazilian Federal District, BA=Bahia state, GO=Goiás state, MT=Mato Grosso state, MS=Mato Grosso do Sul state, MG=Minas Gerais state, PE=Pernambuco state, RJ=Rio de Janeiro state, SP=São Paulo state. The colour and size of the circles indicates the number of new genomes generated in this study (black=DENV1, white=DENV2). **B**. Weekly notified dengue cases normalized per 100K individuals per region in 2015-2020 (until EW06). Epidemic curves are coloured according to geographical macro region: SE=Southeast, NE=Northeast, MW=Midwest, N=North, S=South. **C-E**. Time series of weekly reported cases normalized per 100K individuals and daily mosquito-viral suitability measure (index P) in the three macro regions for which new sequences were generated: MW=Midwest (C), NE=Northeast (D), and SE=Southeast (E). The index P of each region is obtained using average climatic data for the three largest urban centers in each region. Purple bars highlight the dates of sample collection of the genomes generated here. (**B-E**) Incidence (cases per 100K population) is presented in log10 for visual purposes.

Weekly reported incidence (cases normalized per 100K individuals) notified between 2015 and 2020 was aggregated into five Brazilian macro regions: North (N), South (S), Midwest (MW), Northeast (NE), and Southeast (SE) (Figure 1 panel B). Epidemic curves revealed three major outbreaks in all regions during early 2015, 2016, and mid-2019, with the SE region playing such a prominent role that its reported incidence was close to the overall incidence for Brazil (Figure 1 panel B, Figure S2). For the macro regions with new viral sequences (MW, SE, NE), we estimated transmission potential using a mosquito-viral suitability index. Since it is not possible to obtain representative temperature and humidity time series for the macro regions, we instead used climatic series from the three largest urban centers of each region. While the latter neglects climate variation within a macro region (Figure S3), the seasonal timing of reported cases was still well captured by the urban suitability indices (Figure 1 panels C–E). The years 2015, 2016, and 2019 did not show particular increases in suitability, suggesting that the corresponding larger epidemics were not driven by particular climatic trends^29^ but rather by factors not accounted for by the index (e.g. sociodemographic, lack of herd immunity, change in circulating serotype / genotype, size of the mosquito population, etc). There was also a clear decrease in reported cases across the regions during the aftermath of the Zika virus epidemic (2017-2018), a phenomenon also reported elsewhere. ^29–31^ In contrast, reporting of other arboviruses (e.g. chikungunya, Figure S4) presented increases in incidence during the same period. As such, we found no support for either climate driven reductions in transmission potential or arboviral reporting saturation that could explain the drop in DENV reporting between 2017 and 2018, suggesting that local herd immunity (sero-specific or induced by Zika infection) could have played a role. Finally, apart from a few exceptions, the dates of the new sequences matched time periods of both high suitability and case reporting for all regions and are thus representative of epidemic periods (Figure 1 panels C—E).

Between 2015 and 2019, a total of 3,180 deaths attributed to DENV were reported in Brazil. We found a clear seasonal signal in weekly reported deaths that matched the seasonality of weekly reported cases and suitability for transmission (Figure S5). The year 2019 has been widely reported as experiencing a substantial increase in both the number of cases and deaths. Accordingly, we found the MW and NE had an increase in the absolute number of deaths in 2019 compared to previous years (Figure S5 panels B and D). However, when the weekly (crude) case fatality rate (CFR=deaths/cases) was calculated there was no increase in the CFR during 2019 for any of the regions (Figure S5 panel C, E, G). When aggregating the CFR between 2015 and 2019, we found MW to have a higher CFR at 0·00084 (1·78×10^-05^ – 2·01×10^-03^ 95% range) compared to 0·0006 for NE (0-0·0030) and 0·00037 for SE (0-0·0010) (only the MW versus SE and NE comparisons were statistically different using a Wilcoxon test; p-values 5·76e-09 and 3·775e-05, respectively – Figure S5).

Dengue serotypes universally present temporal dynamics with an oscillatory behaviour characterized by recurrent peak prevalence of each serotype every 8-11 years.^32^ Due to limitations in local testing capacity, inferring the relative prevalence of DENV 1-4 with high spatio-temporal resolution in Brazil is often difficult. Our limited data (Table S3) suggests that between 2015 and 2016 DENV1 was the dominant serotype in the MW, NE, and SE regions (Figure S6). Replacement in dominance by DENV2 took place in MW during 2017 and in the following years in SE. Throughout the study period, DENV1 was the dominant serotype in the NE region.

### DENV1 phylodynamics in Brazil, 2015 to 2019

To explore the phylodynamics of DENV1, we combined our 57 newly generated sequences to DENV1-V genomes available on GenBank (n=501). Phylogenetic analysis revealed that the novel isolates were organized into three distinct clades, named hereafter as clades I, II, III (Figure 2 panel A). Clade I appeared to have been replaced by clades II-III during 2019. Hence, the recent epidemic was characterized by the co-circulation of two viral lineages, which suggests that non-virological factors (e.g. susceptibility levels in the human population) were likely responsible for the expansion of DENV1 in 2019.

**Figure 2.**
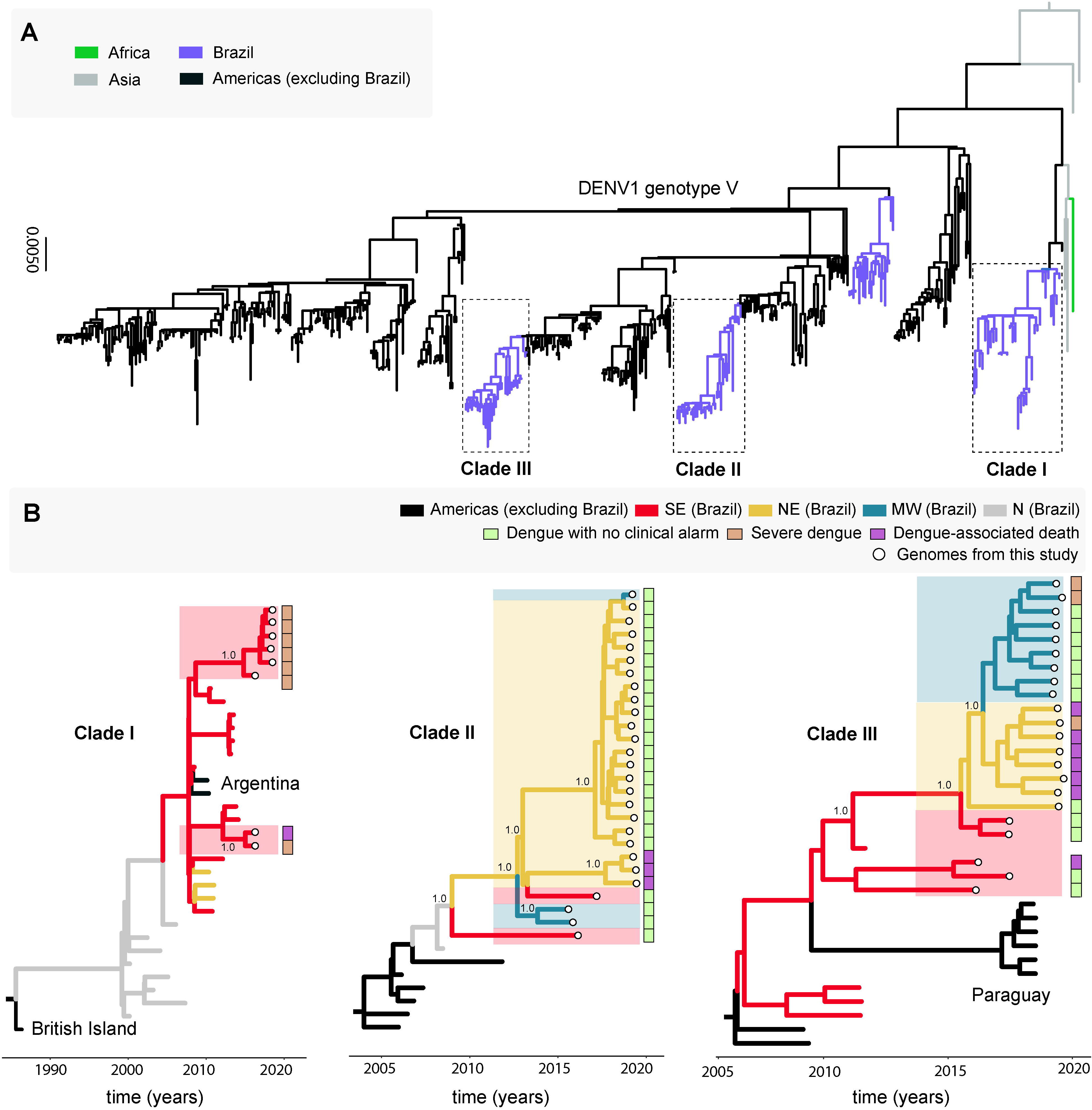
Phylogenetic analysis of DENV1-V in Brazil. **A**. Maximum likelihood (ML) phylogenetic analysis of 57 complete genome sequences from DENV1 generated in this study plus 501 publicly available sequences from GenBank. The scale bar is in units of nucleotide substitutions per site (s/s) and the tree is mid-pointed rooted. Colours represent different sampling locations. **B**. Time-scaled phylogeographic tree of Clade I (including eight new sequences plus 25 GenBank sequences), Clade II (including 27 new sequences plus seven GenBank sequences), and Clade III (including 22 new sequences plus 12 GenBank sequences). Colours represent different sampling locations (SE Brazil=Brazilian Southeast, NE Brazil=Brazilian Northeast, MW=Brazilian Midwest, N Brazil=Brazilian North). Tip circles represent the genome sequences generated in this study; coloured sidebars represent the dengue clinical classification for each sequenced sample.

To investigate the evolution of clades I-III in more detail, we used smaller data sets (n=33, 34, 34, respectively) derived from these clades individually. An analysis of substitution rate constancy using TempEst revealed a strong correlation between the sampling time and the root-to-tip divergence in all three data sets (Figure S7), allowing the use of molecular clock models to infer evolutionary parameters. Phylogeographic analyses of clade I (Figure 2 panel B) clustered the new sequences into a single well-supported monophyletic sub-clade (posterior probability support, PPS=1·0), including isolates sampled between 2000 and 2018. New sequences in this clade were mainly from severe dengue cases registered in the SE region. The tMRCA of all Brazilian sequences was estimated to be between June 1998 to February 2000, and this common ancestor likely originated in the N region (PPS=1·0), after a dispersion of a strain from the British Virgin Islands (PPS=1·0). Viruses from this clade spread from the N region towards SE and then to NE, as indicated by isolates from Pernambuco (PE) state (represented by JX669461, JX669465, and JX669464). The tMRCA of all isolates from SE and NE in this clade was estimated to be between May 2006 to February 2008.

Similarly, an analysis of clade II (Figure 2 panel B) revealed a single well-supported monophyletic group (PPS=10), including isolates from SE, MW, and NE regions sampled between 2015 and 2019. The majority of the new sequences were from mild dengue cases, although three isolates were recovered from fatal cases in NE. The tMRCA of this group was dated to between August 2007 and May 2010, with a likely origin in the N region. However, as the PPS value was low (0·39) the place of origin remains uncertain. After its introduction into the N region, viruses from this lineage appear to have moved towards SE, MW, and NE. Notably, the clade appears to have persisted locally after its introduction in the NE region between July 2011 to June 2014, and from there a second introduction into MW may have occurred, as suggested by a single isolate (OPAS134) sampled in 2019.

Finally, clade III (Figure 2 panel B) also formed a single supported monophyletic group (PPS=0·82) that contained sequences from SE, NE, and MW regions sampled between 2011 and 2019. Among our new isolates, six were from fatal cases reported in NE. We estimated the age of this sub-clade to be between October 2009 to August 2011, with a most likely origin in the SE region (PPS=0·99). Since its introduction, the clade has circulated in SE, from where it has later dispersed to Paraguay. The SE region has also seemingly seeded outbreaks into the NE (PPS=0·88) between February 2015 and September 2017, and subsequently towards the MW between June 2016 to April 2018.

### DENV2 phylodynamics in Brazil, 2016 to 2019

To explore the phylodynamics of DENV2 between 2016 and 2019, we performed a phylogenetic analysis of the 170 newly generated sequences plus 450 complete genome sequences of DENV2-III available on GenBank (Figure 3). This analysis revealed four different clades (termed hereafter BR-1 to BR-4 clades).^33^ Notably, BR-1 contained Brazilian sequences sampled from 1990-2000 BR-2 from 2000-2006, and BR-3 from 2006-2019. The latter included six of our new isolates (3·5%, 6/170) collected during previous outbreaks in 2008 (GO=3) and 2016 (SP=3). Finally, BR-4 contained the other 164 new sequences (96·5%, 164/170), sampled between 2016 and 2019. This phylogenetic pattern suggests that between 2016 and 2019 DENV2 circulated in these Brazilian regions with a succession of different viral clades from to BR-3 and then BR-4, with older clades (BR-1, BR-2) either not being sampled in the most recent time-points or having experienced local extinction. These phylogenetic results are consistent with a recent study highlighting the role of the Caribbean region in the spread of the BR-4 clade into Brazil^29,33^ (Figure 3 panel A).

**Figure 3.**
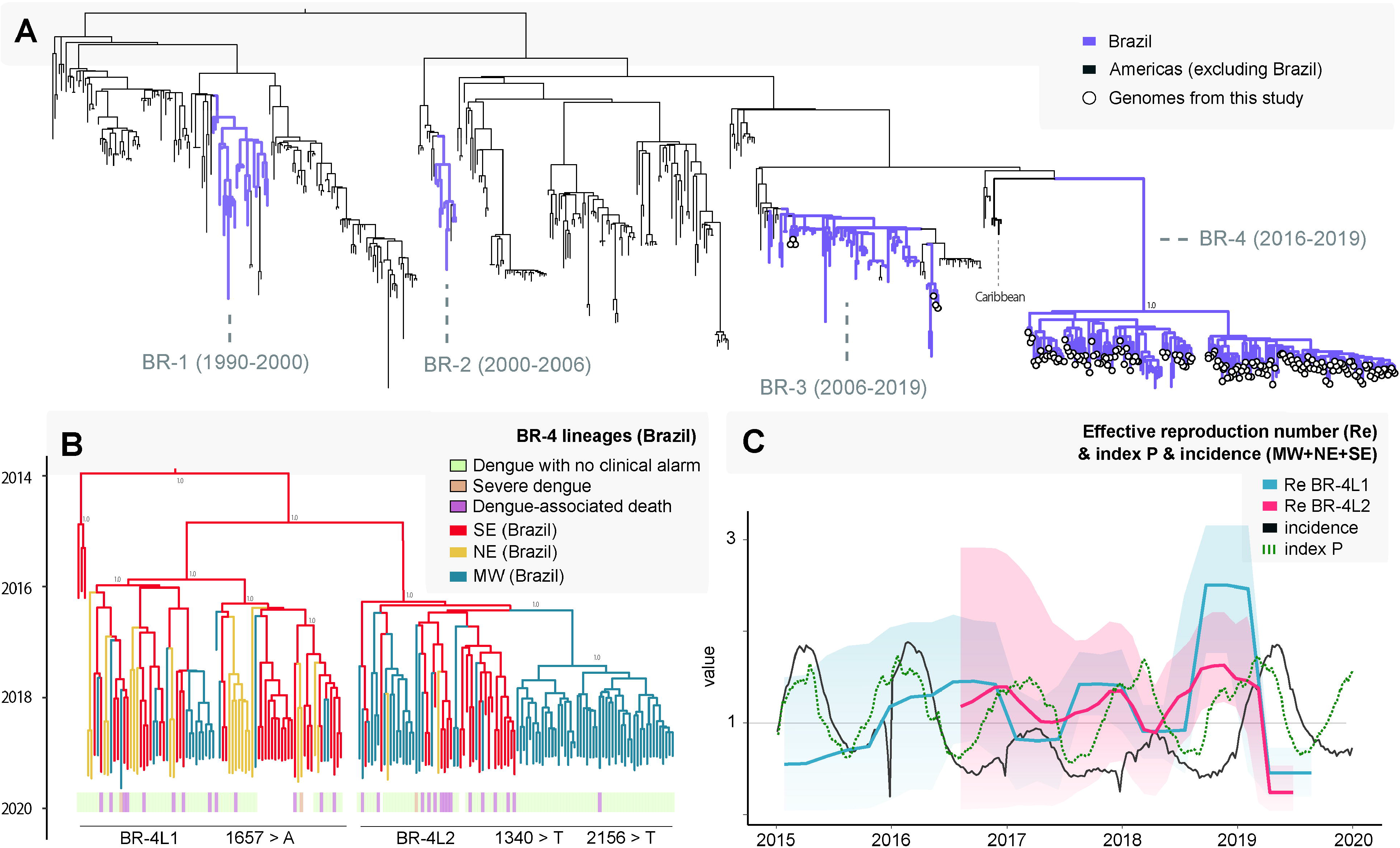
Phylogenetic analysis of DENV2-III in Brazil. **A**. A maximum likelihood tree was inferred using 170 complete genome sequences from DENV2 generated in this study and 450 sequences publicly available, retrieved from GenBank. The scale bar is in units of nucleotide substitutions per site (s/s) and mid-point rooted. Tip circles represent the genome sequences generated in this study. **B**. Time-scaled phylogeographic DENV2 BR-4 tree, including 164 new DENV2 sequences generated here and 17 publicly available data from the 2019 outbreak in Brazil.^33^ Sequences are coloured according to sampling location. Sidebars represent the dengue clinical classification for each sequenced sample. **C**. Temporal fluctuation of the effective reproduction number (R_e_) of the for DENV2 BR-4L1 (blue) and BR-4L2 (magenta) estimated using the Bayesian birth-death approach. Black line is the total weekly incidence of dengue between 2015 and 2020 (until EW06), and the dotted green line is the index P (incidence is summed and index P is averaged over the three macro regions for which new sequences were generated: MW=Midwest, NE=Northeast, and SE=Southeast). Incidence (cases per 100K population) is presented in log10 for visual purposes.

Given the substantial number of novel sequences, we examined the BR-4 clade in more detail using a molecular clock approach (Figure 3 panel B) (and a linear regression of root-to-tip genetic distance against sampling date revealed sufficient temporal signal, r^2^=0·60; Figure S8). The BR-4 clade (PPS=1·0) included 181 isolates from the SE, NE, and MW regions, 92·3% (167/181) of which were sampled during the 2019 outbreak and 7·7% (14/181) sampled between 2016 and 2018. Of these, 133 were from dengue cases without severe disease, while three and 28 isolates were recovered from severe and fatal dengue cases, respectively. We identified two distinct BR-4 lineages circulating between 2017 and 2019, which we termed BR-4L1 and BR-4L2 (Figure 3 panel B). Both lineages contained sequences from the NE, SE, and MW regions. The tMRCAs of BR-4L1 and BR-4L2 were dated to between September 2014 and June 2016 and March 2015 to November 2016, respectively. BR-4L2 contained a monophyletic cluster of isolates from MW sampled between 2018 and 2019. We also observed that the other isolates from NE and MW were intermixed throughout both lineages, indicating that multiple introductions of DENV2 over time. Similar to recent studies,^33^ we found the tMRCA of BR-4 to be between November 2013 and May 2015, likely in SE (PPS=0·99), from where the virus dispersed towards NE and MW regions. MG state, located in SE, seems to have played an important role as source location as sequences from this region (from 2016) fell close to the root of the clade (Figure 3 panel B).

We identified 34 single nucleotide variants between the BR-4L1 and BR-4L2 lineages, of which only three resulted in amino acid substitutions. Isolates of the BR-4L1 acquired one unique amino acid substitution V553I (NS5 protein), while only a few isolates of this lineage had a second amino acid substitution K719I (NS5 protein). All isolates of BR-4L2 acquired two unique amino acid substitutions A447V and K719I (in the ENV and NS5 proteins, respectively) (Table S4).

We used a BDSKY model to tentatively estimate the R_e_ of BR-4L1 and BR-4L2 (Figure 3 panel C). This provided evidence for three significant seasonal oscillations in R_e_ (although with wide credible intervals), consistent but generally preceding the time windows of reported outbreaks between 2016 and 2019 (Figure 1 panels C-E). Mosquito-viral suitability presented the same general patterns, but the timing of its oscillations was in between that of R_e_ and incidence (Figure 3 panel C). In general, our estimates of R_e_ for both lineages peaked at the end of each year, decreasing and remaining below 1 temporarily at the start of each following year (although again note the credible intervals). Notably, the time period with the largest R_e_ for both lineages at the end of 2018 and preceding the large epidemic of 2019 (in excess of 2·5 for BR-4L1 and 1·5 for BR-4L2) did not coincide with similar increases in suitability.

## DISCUSSION

More than 16 million cases of dengue disease have been notified since the early 1980s in Brazil.^7,12^ Previous studies have explored the evolution of DENV1-4 in the Americas, mainly focusing on a restricted range of countries using partial genome sequences.^34-37^ To obtain a better understanding of DENV evolution in Brazil we generated 227 new complete genome sequences of DENV1 and DENV2 through portable sequencing. Over three quarters of the new sequences were processed and analyzed during a Nanopore-based genome sequencing training and surveillance program that took place in Belo Horizonte, MG state, in 2019. The new 227 sequences generated correspond to 55% (57/104) and 60% (170/285) of DENV1-V and DENV2-III, respectively, of Brazilian complete genomes that are currently available in public databases (Figure S9). This highlights the large contribution of this study, but also the current shortage of complete genome data for these serotypes. There is clearly a need for continued funding for genomic surveillance, which, as described here, can contribute to a better understanding of the introduction, spread, and persistence of dengue viruses in Brazil.

Time series of reported cases between 2015 and 2019 showed the typical yearly seasonal patterns of dengue transmission. Reporting was low in 2017 and 2018, coinciding with the post-epidemic period of the Zika virus in Brazil. Such trends have also been reported elsewhere,^29,30^ and are speculated to be driven by transient cross-protection from exposure to Zika and/or temporary saturation or changes in surveillance.^38^ When comparing to reported cases of chikungunya virus and estimated mosquito-viral suitability in the same period, we found no evidence of changes in capacity for arboviral surveillance or climate driven low transmission potential, in favor of other mechanisms playing a role (e.g. Zika cross-immunity). In contrast, there were three particularly large DENV epidemics: in 2015 and 2016 when DENV1 was dominant across all regions, and in 2019 when DENV1 was dominant in the NE but DENV2 had become dominant in the MW and SE. Due to the increased likelihood of secondary infections, serotype replacement is often associated with measurable changes in the clinical spectrum of reported cases, with increases in both disease severity and number of deaths.^9,37,39^ While we describe an increase in absolute case and death numbers in 2019, the emergence of DENV2 in the SE and MW was not associated with a significant increase in the case fatality rate compared to previous years.

Our newly generated DENV1 sequences were classified as genotype V and formed three distinct clades (I-III). This corroborates previous reports suggesting that such clades remain persistent and are responsible for the latest DENV1 outbreaks in Brazil.^35,36,39,40^ Within clade I only eight of the new sequences were sampled between 2016-2018, while many isolates preceded 2015. The shortage of genomic data in the intermediate years severely hampered our capacity to draw further conclusions, such as the possibility of a temporary lineage replacement event. In contrast, most of the new sequences within clades II-III were sampled in 2019, supporting the co-circulation of two DENV1 lineages in recent epidemics – a phenomenon often described in other DENV epidemics.^35,36,39,40^ Isolates from the three clades have been identified in both SE and NE regions; but the most recent isolates from clade I appear only in the SE, while isolates from clades II-III appear in the NE and MW. While this suggests some structure in spatio-temporal circulation, we cannot ascertain if it is due to a bias resulting from limited sampling. Time estimates the tMRCA of the new Brazilian isolates (2015-2019) were between May 2006 and February 2008 for clade I, between August 2007 and May 2010 for clade II, and between October 2009 to August 2011 for clade III. Such large ranges are compatible with recent estimates showing also large uncertainty^29^ likely due to lack of genomic data during this period, reinforcing the need for a more effective surveillance in Brazil.

All new DENV2 complete genome sequences belong to genotype III, which has also been found in previous epidemics in Brazil.^33,41^ Our results are also in line with reports of three different lineages causing outbreaks in Brazil since 1990,^40,41^ and support studies describing a recent introduction of DENV2-III.^33^ We found that isolates from this genotype were grouped in four different clades (BR-1 to BR-4) with apparent replacement over time. Specifically, the oldest lineage BR-1, including isolates from 1990-2000, was replaced by BR-2 comprising sequences from 2000-2006, subsequently replaced by BR-3 containing isolates from 2006-2019. Finally, BR-3 was replaced by BR-4, containing sequences sampled between 2016-2019, some closely related to Caribbean isolates sampled in 2005. In a similar manner to DENV1, DENV2 BR-3 and BR-4 isolates from 2019 demonstrated the co-circulation of at least two different lineages in recent years.^33^ The relationship to Caribbean sequences suggests a possible origin in this region, although there is a large temporal gap between the sampling of the Caribbean sequences from 2005 and the early Brazilian sequences from 2016, again highlighting the need for more genomic surveillance in Brazil. The tMRCAs of BR-4L1 and BR-4L2 were dated September 2014 to June 2016 and March 2015 to November 2016, respectively, which may coincide with the emergence and spread of the Zika^42^ and chikungunya^43^ viruses, and a high incidence of dengue in Brazil. Six years after introduction, the lineages continue to circulate in the SE region, and were present in the most recent large epidemic of 2019. From SE, dispersion was towards NE and MW, with multiple independent introductions identified.

Analysis of the 181 isolates from three macro regions (NE, MW, SE) allowed us to estimate the emergence of DENV2 BR-4 in SE between November 2012 and May 2015, supporting previous reports.^29,33^ After its introduction, BR-4 has circulated as two distinct lineages (BR-4L1 and BR-4L2). We observed three single nucleotide variants among the lineages that resulted in amino acid substitutions: V553I and A447V were identified in the BR-4L1 and BR-4L2, respectively, while K719I was identified in both lineages. A447V and V553I, mapped to ENV and NS5, respectively, appear to be conservative changes due to the interchangeable character for the respective amino acids.^44^ In contrast, K719I in the NS5 protein has changed from a negatively charged to an aliphatic amino acid.^44^ Further studies are required to elucidate the impact of these variants on structure and function of the associated proteins, and any potential role in both viral pathogenesis and fitness.

Our retrospective reconstruction of the recent transmission history of DENV1 and DENV2 revealed that the SE and N regions of Brazil were key to the dispersal in Brazil. This supports previous studies that have highlighted both regions as important hubs for introduction and dispersion in the country not only of DENV,^7-10,35,39,41^ but also for the yellow fever virus.^45^ By combining genetic and epidemiological models we show that the establishment and the co-circulation of the DENV2 BR-4L1 and BR-4L2 in several Brazilian regions occurred during a time window of sustained transmission potential measured by estimates of R_e_ and mosquito-viral suitability. These results are consistent with sufficient ecological suitability for the virus’s main vectors *(Aedes* spp.) and insufficient population-level herd-immunity, supporting the expectation of continuing endemic circulation of these dengue viruses in Brazil.

The new genomic data presented here has been generated using portable sequencing tools in a field surveillance initiative (ZiBRA-2 project) and a genomic surveillance training program. We present a range of research outputs describing the recent history and genomic epidemiology of DENV1 and DENV2 in Brazil, corroborating previous studies and greatly increasing the number of public viral genome sequences available for analysis. Several of our outputs identify gaps in existing genomic data, which curtail definite conclusions on key points of the recent history of DENV in Brazil. Importantly, epidemiological and genomic data was analysed in real-time during the training program and subsequently during online sessions, and the research outputs presented here had a significant contribution of the participants attending the program in August 2019. We call for continued funding of similar field and classroom genomic surveillance initiatives to those described in this article. These have the potential to build local capacity in the field of genomic surveillance and in doing so, advancing our understanding on the population-biology of circulating arboviruses and those yet to emerge.

## Data Availability

Newly generated DENV1 and DENV2 sequences have been deposited in GenBank and accession numbers will be provided upon final submission. All supplementary files can be found at: https://www.zibra2project.org/data/.

## CONTRIBUTORS

Conception and design: TERA, MG, VF, JL, and LCJA; **Data collection:** TERA, MG, VF, LHFD, MAAO, VLS, ALSM, GMC, RHS, ECO, JACJ, FCMI, ANBF, ALA, RJ, CFCA, JMR, RFCS, JAS, NFOM, PL, LCVF, SKH, AM, FKB, CCV, RVC, ELLA, SFOT, AAF, FLLC, PLS, FBN, GGCL; MCSUZ, FMCS, CCMG, ZDCFG, SNS, LAP, ANM, FMP, JJFM, ACMSJ, MML, RMRN, AGN, VACA, DBR, WKO, ACM, VP, Latin American Genomic Surveillance Arboviral Network, ECH, TdO, JL, and LCJA; **Investigations:** TERA, MG, VF, LHFD, MAAO, VLS, ALSM, GMC, RHS, ECO, JACJ, FCMI, ANBF, ALA, RJ, CFCA, JMR, RFCS, JAS, NFOM, PL, LCVF, SKH, AM, FKB, CCV, RVC, ELLA, SFOT, AAF, FLLC, PLS, FBN, GGCL; MCSUZ, FMCS, CCMG, ZDCFG, SNS, LAP, ANM, FMP, JJFM, ACMSJ, MML, RMRN, AGN, VACA, DBR, WKO, ACM, VP, Latin American Genomic Surveillance Arboviral Network, ECH, TdO, JL, and LCJA; **Data Analysis:** TERA, MG, VF, LHFD, MAAO, VLS, ALSM, GMC, RHS, ECO, JACJ, FCMI, ANBF, ALA, RJ, CFCA, JMR, RFCS, JAS, NFOM, PL, LCVF, SKH, AM, FKB, CCV, RVC, ELLA, SFOT, AAF, FLLC, PLS, FBN, GGCL; MCSUZ, FMCS, CCMG, ZDCFG, SNS, LAP, ANM, FMP, JJFM, ACMSJ, MML, RMRN, AGN, VACA, DBR, WKO, ACM, VP, Latin American Genomic Surveillance Arboviral Network, ECH, TdO, JL, and LCJA; **Writing – Original:** TERA, MG, VF, JX, ASA, FKB, SFOT, FBN, ECH, TdO, JL, and LCJA; Draft Preparation: TERA, MG, VF, JX, ASA, FKB, SFOT, FBN, ECH, TdO, JL, and LCJA; Revision: TERA, MG, VF, JX, ASA, FKB, SFOT, FBN, ECH, TdO, JL, and LCJA; **Resources:** LCJA, LHFD, MAAO, VLS, ALSM, RHS, ECO, JACJ, ANBF, SKH, AM, ALA, RJ, CFCA, JMR, RFCS, JAS, NFOM, PL, LCVF, RVC, ELLA, DBR, WKO, and ACM.

## DATA SHARING

Newly generated DENV1 and DENV2 sequences have been deposited in GenBank and accession numbers will be provided upon final submission. All data including alignments, tree files as well as epidemiological data can be found at: https://www.zibra2project.org/data/.

## COURSE MATERIAL

Information about the genomic surveillance training program can be found in Supplementary Text File 1.

## ACKNOWLEDGMENTS

The authors thank all the members of the Latin American Genomic Surveillance Arboviral Network (a full list of the participants can be found in the Supplementary Material – Table S5). The authors also thank all personnel from the Health Surveillance System of the Brazilian Federal District and from the states of Bahia, Goiás, Mato Grosso, Mato Grosso do Sul, Minas Gerais, Pernambuco, Rio de Janeiro, and from Blood Center of Ribeirao Preto (Sâo Paulo, Brazil) that helped with sampling and epidemiological data collection. MG is supported by Fundaçâo de Amparo a Pesquisa do Estado do Rio de Janeiro (FAPERJ). VF and TdO are supported by the South African Medical Research Council (MRC-RFA-UFSP-01-2013/UKZN HIVEPI) and the NIH H3AbioNet network, which is an initiative of the Human Health and Heredity in Africa Consortium (H3Africa). ECH is supported by an Australian Research Council Australian Laureate Fellowship (FL170100022). JL is supported by a lectureship from the Department of Zoology, University of Oxford. JX, VF and FCMI is supported by the Coordenaçâo de Aperfeicoamento de Pessoal de Nível Superior – Brasil (CAPES) – Finance Code 001. FI is also supported by Fundaçâo de Amparo à Pesquisa do Estado de Minas Gerais (FAPEMIG). ASA has a scholarship from ZIKA – Announcement MCTIC/FNDCT-CNPq/MEC-CAPES/MS-Decit /No. 14/2016 – Prevention and Fight against Zika Virus.

## SUPPLEMENTARY FILES

All supplementary files can be found at: https://www.zibra2project.org/data/.

Supplementary File 1 – Table with estimated index P for Brasília, Goânia, Campo Grande, Sâo Paulo, Rio de Janeiro, Belo Horizonte, Salvador, Recife, Fortaleza (CSV).

Supplementary File 2 – Reported cases and deaths in MW, NE, SE (CSV).

Supplementary File 3 – Alignments and Phylogenetic trees (.fasta and .tree files).

Supplementary File 4 – List of authors indexing.

Supplementary File 5 – Supplementary Appendix.

